# KNOWLEDGE, ATTITUDE, AND PERCEPTION OF MOTHERS OF UNDER-FIVE CHILDREN TOWARD CHILDHOOD ROUTINE IMMUNIZATION PROGRAMME IN ABUJA MUNICIPAL AREA COUNCIL, NIGERIA

**DOI:** 10.1101/2025.08.14.25332517

**Authors:** Bukola Sarah Ogunleye, Sylvester Sesugh Shakpande

**Affiliations:** Ahmadu Bello University, Zaria, Nigeria

**Keywords:** Childhood immunization, knowledge, attitudes, perceptions, mothers, primary healthcare, Abuja Municipal Area Council (AMAC), Nigeria, chi-square test

## Abstract

**Background:** Vaccine-preventable diseases (VPDs) remain a major public health challenge among children under five in Nigeria, despite sustained government efforts to provide access to essential vaccines. This study assessed the knowledge, attitudes, and perceptions of mothers with under-five children regarding routine childhood immunization at primary healthcare centers in Abuja Municipal Area Council (AMAC), Federal Capital Territory, Nigeria.

**Methods:** A cross-sectional study was conducted using a multi-stage sampling technique to select 281 mothers of under-five children. Data were collected using structured questionnaires covering sociodemographic characteristics and knowledge, attitudes, and perceptions (KAP) regarding routine immunization. Descriptive statistics were used to summarize the data, and chi-square tests were applied to identify associations between sociodemographic variables and KAP outcomes at a significance level of p < 0.05.

**Results:** Most respondents (94.7%) recognized the importance of routine immunization in preventing VPDs. However, 37.4% lacked awareness of specific vaccines required, and 13.2% considered vaccines potentially harmful. While 97.2% reported adhering to the immunization schedule, uptake of supplementary vaccines was low (17.8%) due to financial constraints. Overall perceptions were positive, with 97.5% viewing vaccines as safe, though sociocultural factors, particularly religion, influenced these perceptions. Statistically significant associations were observed between household income and both knowledge (χ = 25.930, p < 0.001) and perception (χ = 10.230, p = 0.010), with mothers from lower-income households demonstrating better knowledge and more favorable perceptions. Higher educational attainment was also significantly associated with better knowledge (χ = 32.250, p < 0.001). No significant associations were found for age, marital status, employment status, or ethnicity (p > 0.05).

**Conclusion:** Although mothers in AMAC generally have positive attitudes and perceptions toward routine immunization, critical knowledge gaps persist. Socioeconomic and educational factors significantly influence maternal understanding and acceptance. Targeted health education and economic support initiatives are essential to bridge knowledge gaps and enhance vaccine uptake in this population.

## Introduction

Vaccine-preventable diseases (VPDs) continue to be a major cause of morbidity and mortality among children under five years old, particularly in low- and middle-income countries (World Health Organization, 2013). The World Health Organization (WHO) estimates that immunization prevents 4 million deaths annually from diseases such as diphtheria, tetanus, pertussis, influenza, and measles (Centers for Disease Control and Prevention, 2023). The vaccination of a larger population against such diseases results in the generation of “herd immunity”. This helps confer protection to the community at large, particularly the unvaccinated. To achieve herd immunity, a target of 90% to 95% immunization coverage is required (Dassarma *et al.,* 2022). Although about one-third of child deaths can be prevented among the under-five, sadly, a child still dies from a vaccine-preventable disease every 20 seconds (Adefolalu *et al.,* 2019). Records show that about 13.5 million children missed out globally on basic childhood vaccines in 2022 (World Health Organisation, 2024), with Africa and Asia bearing the brunt, accounting for over 70% of the world’s unvaccinated children (Adefolalu *et al.,* 2019). Studies show that one in five children residing in Africa has not received all the necessary vaccinations based on WHO guidelines.

Over 30 million under-five children have VPD every year, while over 500,000 children die annually, leaving Africa responsible for about 58% of VPD-related deaths globally (World Health Organization, 2024). Despite global progress, sub-Saharan Africa continues to experience low immunization coverage, contributing significantly to child mortality. According to a study conducted by Mboussou *et al.,* (2024), African vaccine coverage for the first and third doses of the diphtheria-tetanus-pertussis-containing vaccine (DTP1 and DTP3) was estimated at 80% and 72%, respectively, while the first dose of the measles-containing vaccine (MCV1) was 69% in 2022. It is important to note that only 13 of the 47 countries (28%) in Africa were able to achieve global vaccine coverage of 90% or above with DTP3 in 2022. 1 in 5 African children did not complete their routine basic vaccines, leaving over 30 million children under five vulnerable to vaccine-preventable diseases (VPDs) annually. Thus, Africa accounts for about 58% of global VPD-related deaths (World Health Organization, 2024).

In Nigeria, the Expanded Programme on Immunization (EPI) was introduced in 1979 to reduce the burden of childhood diseases (WHO & UNICEF, Geneva: WHO, 2023). Routine immunization services are offered free of charge at public health facilities. However, it is disheartening to note that immunization coverage in Nigeria is below GVAP goals, putting a significant number of children’s lives at risk. According to the report of the Multiple Indicator Cluster Survey (MICS) and National Immunization Coverage Survey (NICS), in 2021, only 57% of children between 12-23 months of age in Nigeria completed their three doses of pentavalent vaccine. In addition, 21% of Nigerian children who received penta-1 dose did not receive penta-3 dose. Pentavalent vaccine monitoring is important because it serves as an indicator of routine childhood vaccine monitoring. Furthermore, 46% of the children received incomplete routine vaccines, while 18% did not receive any dose, indicating that 64% of children aged 12-23 months did not receive all the recommended routine immunizations. Vaccine benefits are fully realized when children take all the recommended doses of the vaccine promptly. Overall, only 36% of children aged 12-23 months received all recommended vaccines in Nigeria.

Immunization coverage varies across the nation. According to the MICS and NICS survey 2021, only two states, namely Ebonyi and Enugu, have recorded penta-3 vaccine coverage above 90%, while the Federal Capital Territory of Nigeria, Abuja, was below the target goal. The estimated Abuja routine childhood vaccine coverage is 80%, while the zero-dose estimate is 4%.

Uptake of these vaccines remains suboptimal in many areas, including the Federal Capital Territory. Several studies have linked low immunization coverage to poor maternal knowledge, negative attitudes, and sociocultural beliefs, which are known to influence healthcare decisions for young children (Adedire *et al.,* 2021).

Mothers, as primary caregivers, play a critical role in the health-seeking behavior and immunization status of their children (Adedire *et al.,* 2021). Their knowledge and perceptions regarding vaccines can significantly influence whether or not a child completes the recommended immunization schedule. Understanding the knowledge, attitudes, and perceptions (KAP) of mothers toward routine immunization is therefore essential to identify barriers and inform effective interventions aimed at increasing vaccine uptake (Shahzadi *et al.,* 2022).

This study was conducted to assess the knowledge, attitudes, and perceptions of mothers of under-five children attending primary healthcare centers in Abuja Municipal Area Council (AMAC), Nigeria, towards routine childhood immunization. It also seeks to identify the key factors influencing these perspectives to support more effective immunization strategies and interventions. The findings are expected to guide public health policy and community-based health education strategies targeted at improving immunization coverage.

## Methods

### Study Design and Setting

This study employed a descriptive cross-sectional design and was conducted in Abuja Municipal Area Council (AMAC), one of the six area councils in the Federal Capital Territory (FCT), Nigeria. The study focused on mothers of under-five children attending selected primary healthcare (PHC) centers within AMAC.

### Study Population and Sampling Technique

The study population comprised mothers who had at least one child under five years of age and who were attending immunization clinics at selected PHCs during the study period. A multi-stage sampling technique was used to select participants. First, AMAC was purposively selected due to its high population density and availability of PHC services. Then, five PHCs were randomly selected. Finally, systematic random sampling was used to recruit 281 eligible mothers from the selected health facilities.

### Data Collection Instrument and Procedure

Data were collected using a pre-tested, structured, interviewer-administered questionnaire. The questionnaire was designed to gather information on sociodemographic characteristics and assess participants’ knowledge, attitudes, and perceptions regarding routine childhood immunization.

Data collection was done with the participants, and questionnaires were completed face-to-face in either English or local languages, depending on the participant’s preference.

### Data Analysis

Data were entered into Microsoft Excel and analyzed using IBM SPSS Statistics version 23. Descriptive statistics, including frequencies and percentages, were used to summarize demographic characteristics and KAP responses. Chi-square tests were used to explore associations between sociodemographic variables and KAP outcomes. Statistical significance was set at p < 0.05.

### Ethical Considerations

Ethical approval for this study was obtained from the Department of Public Health, Ahmadu Bello University, Zaria, while permission was granted by the Federal Capital Territory Health Research Ethics Committee. Informed consent was obtained from all participants, and confidentiality was maintained throughout the study.

## Result

Table 1 shows the sociodemographic characteristics of the respondents. A total of 281 mothers of under-five children were interviewed across selected primary health care centres in Abuja Municipal Area Council (AMAC), Nigeria. The majority of respondents were between 25 and 34 years of age (52.7%), while 24.9% were aged 35–44 years. A smaller proportion (16.0%) were aged 18–24, and only 6.4% were aged 45 and above. Most of the respondents were married (85.4%), while 9.3% were single, and 5.3% were either divorced or widowed. In terms of educational attainment, 40.2% of the women had completed secondary education, 24.2% had tertiary education, another 24.2% had only primary education, and 11.4% had no formal education. Regarding parity, 42.3% had between 3 to 4 children, 30.6% had 1–2 children, while 27.0% had five or more children.

**Table 1.**
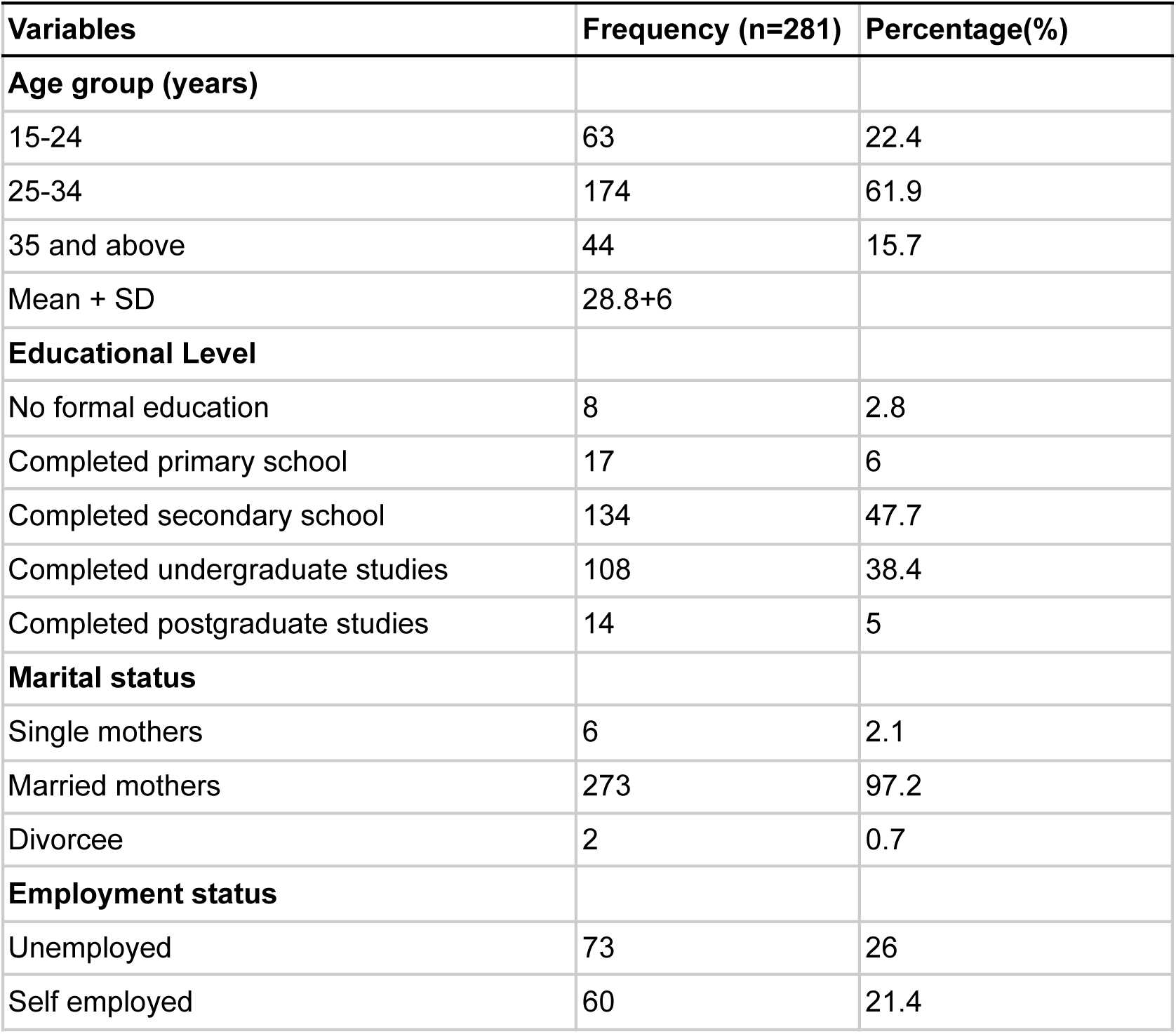
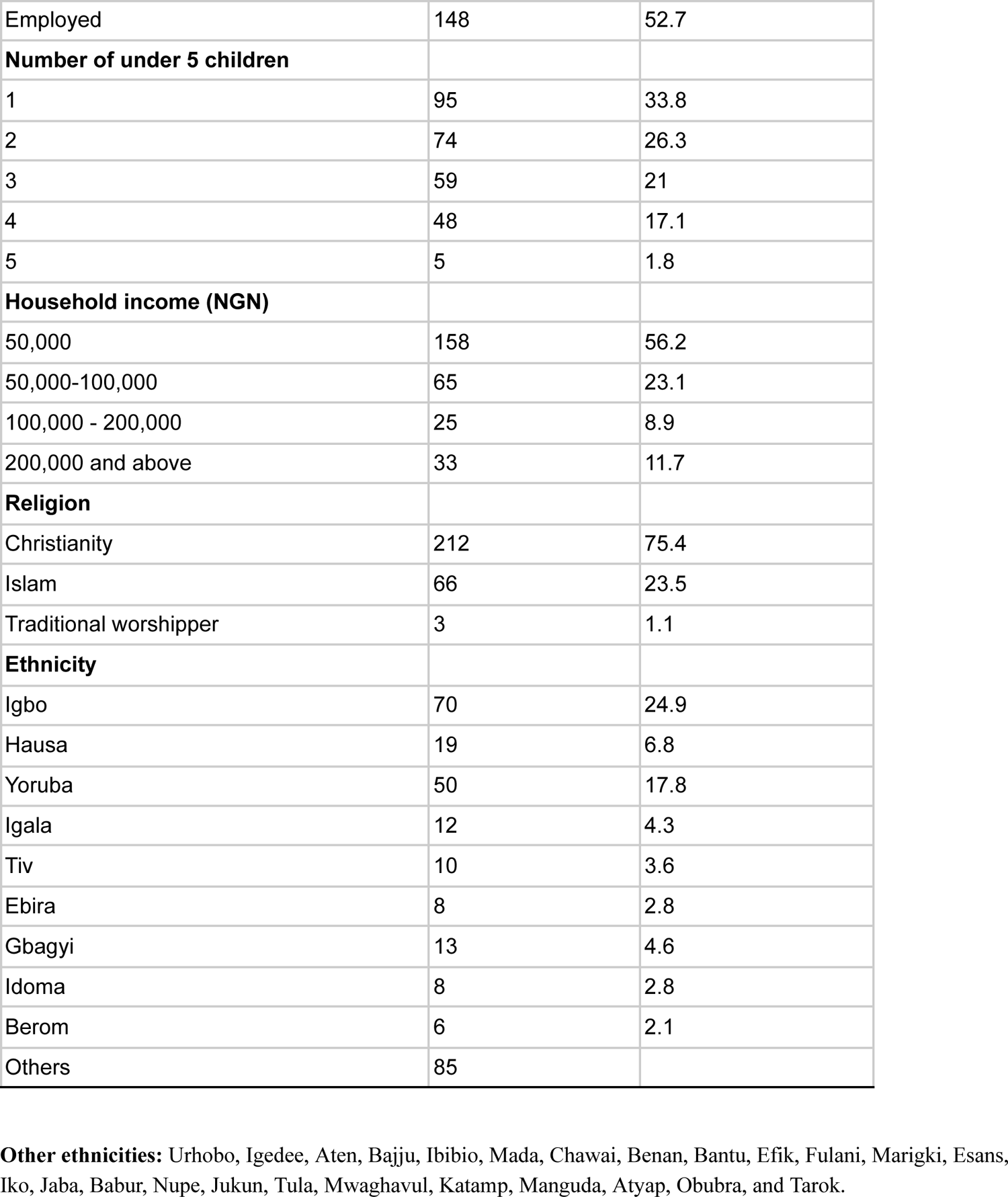
Sociodemographic characteristics of the respondents (n=281)

| Variables | Frequency (n=281) | Percentage(%) |
| --- | --- | --- |
| **Age group (years)** |  |  |
| 15-24 | 63 | 22.4 |
| 25-34 | 174 | 61.9 |
| 35 and above | 44 | 15.7 |
| Mean + SD | 28.8+6 |  |
| **Educational Level** |  |  |
| No formal education | 8 | 2.8 |
| Completed primary school | 17 | 6 |
| Completed secondary school | 134 | 47.7 |
| Completed undergraduate studies | 108 | 38.4 |
| Completed postgraduate studies | 14 | 5 |
| **Marital status** |  |  |
| Single mothers | 6 | 2.1 |
| Married mothers | 273 | 97.2 |
| Divorcee | 2 | 0.7 |
| **Employment status** |  |  |
| Unemployed | 73 | 26 |
| Self employed | 60 | 21.4 |
| Employed | 148 | 52.7 |
| **Number of under 5 children** |  |  |
| 1 | 95 | 33.8 |
| 2 | 74 | 26.3 |
| 3 | 59 | 21 |
| 4 | 48 | 17.1 |
| 5 | 5 | 1.8 |
| **Household income (NGN)** |  |  |
| 50,000 | 158 | 56.2 |
| 50,000-100,000 | 65 | 23.1 |
| 100,000 - 200,000 | 25 | 8.9 |
| 200,000 and above | 33 | 11.7 |
| **Religion** |  |  |
| Christianity | 212 | 75.4 |
| Islam | 66 | 23.5 |
| Traditional worshipper | 3 | 1.1 |
| **Ethnicity** |  |  |
| Igbo | 70 | 24.9 |
| Hausa | 19 | 6.8 |
| Yoruba | 50 | 17.8 |
| Igala | 12 | 4.3 |
| Tiv | 10 | 3.6 |
| Ebira | 8 | 2.8 |
| Gbagyi | 13 | 4.6 |
| Idoma | 8 | 2.8 |
| Berom | 6 | 2.1 |
| Others | 85 |  |

**Other ethnicities:** Urhobo, Igedee, Aten, Bajju, Ibibio, Mada, Chawai, Benan, Bantu, Efik, Fulani, Marigki, Esans, Iko, Jaba, Babur, Nupe, Jukun, Tula, Mwaghavul, Katamp, Manguda, Atyap, Obubra, and Tarok.

### Knowledge of Routine Childhood Immunization

Table 2 shows that the majority of mothers (94.7%) understood the importance of vaccination, though significant knowledge gaps existed regarding vaccine specifics. While 90% correctly identified the birth dose timing, 37.4% couldn’t name specific vaccines, and 35.6% were unaware of common side effects. Chi-square analysis, as observed in Table 3, revealed that education level does significantly impact mothers’ knowledge (χ=32.25, p<0.001), with tertiary-educated mothers demonstrating better understanding than those with only primary education.

**Table 2.** Knowledge of routine childhood immunization among mothers of under-five children in AMAC, Abuja (N = 281)

| Knowledge Item | Correct Response n (%) | Incorrect/ I don't know n (%) |
| --- | --- | --- |
| Recognises vaccine importance | 266 (94.7) | 15 (5.3) |
| Aware of vaccines specific | 176 (62.6) | 105(37.4) |
| Knows vaccines effects | 181 (64.4) | 100 (35.6) |
| Believe vaccines are dangerous | 37 (13.2) | 244 (86.8) |
| Knows the correct timing of first vaccines | 253 (90.0) | 28 (10.0) |

**Table 3.** Relationship between educational attainment and vaccine knowledge scores among participants.

| Education Level | Poor Knowledge n (%) | Good Knowledge n (%) | $\chi^2$ | p-value |
| --- | --- | --- | --- | --- |
| No formal education | 4 (4.9) | 4 (2.0) | 32.250 | <0.001* |
| Primary | 4 (4.9) | 13 (6.5) |  |  |
| Secondary | 51 (63.0) | 83 (41.5) |  |  |
| Undergraduate | 20 (25.0) | 88 (44.0) |  |  |
| Postgraduate | 2 (2.5) | 12 (6.0) |  |  |

### Attitudes Toward Childhood Immunization

Analysis of maternal attitudes (Table 4) revealed strong compliance with vaccination protocols, as evidenced by 273/281 (97.2%) mothers maintaining immunization cards and following the recommended vaccination schedule. However, only 17.8% obtained supplementary vaccines, with 53% citing cost as a significant barrier. Logistic regression found no statistically significant predictors of attitude (all p>0.05), though higher income showed a non-significant trend toward positive attitudes (aOR=3.52, p=0.277) as demonstrated by Table 5.

**Table 4.** Attitudes regarding childhood vaccination among mothers of under-five children (N = 281)

| Attitude Item | Positive Response n (%) | Negative Response n (%) |
| --- | --- | --- |
| Maintains immunisation card | 273 (97.2) | 8 (2.8) |
| Complete required vaccines | 272 (96.8) | 9 (3.2) |
| Follows the immunisation schedule | 273 (97.2) | 8 (2.8) |
| Obtains supplementary vaccines | 50 (17.8) | 231 (82.2) |
| Affected by vaccine costs | 149 (53.0) | 132 (47.0) |
| Mothers who will recommend vaccines to their female friends | 266 (94.7) | 15 (5.3) |

**Table 5.**
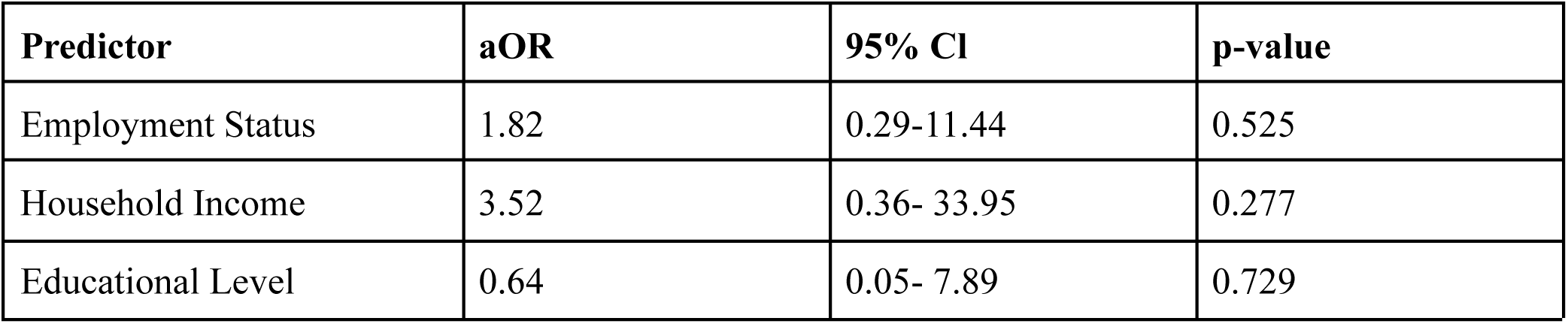
Predictors of maternal attitudes toward childhood immunization: Logistic regression results (N = 281)

| Predictor | aOR | 95% CI | p-value |
| --- | --- | --- | --- |
| Employment Status | 1.82 | 0.29-11.44 | 0.525 |
| Household Income | 3.52 | 0.36- 33.95 | 0.277 |
| Educational Level | 0.64 | 0.05- 7.89 | 0.729 |

### Perceptions of Vaccine Safety and Effectiveness

Table 6.0 revealed that perceptions were predominantly positive, with 97.5% believing in vaccine safety and 92.2% trusting health workers’ advice. However, income significantly influenced perceptions (χ=10.23, p=0.01), with lower-income mothers showing more positive views as observed in Table 7.0. Surprisingly, mothers earning ₦100,000-200,000 had 91% lower odds of positive perception compared to the lowest income group (aOR=0.09, p=0.025).

**Table 6.** Perceptions regarding routine childhood immunization among mothers (N = 281)

6.0 Perceptions regarding routine childhood immunization among mothers (N = 281)
| Perception Item | Positive Perception n (%) | Negative Perception n (%) |
| --- | --- | --- |
| Believe vaccines are effective | 155 (55.2) | 126 (44.8) |
| Thinks government support is adequate | 244 (86.9) | 37 (13.1) |
| Believes vaccines are safe | 274 (97.5) | 7 (2.5) |
| Feels fulfilled vaccinating a child | 275 (97.9) | 6 (2.1) |
| Trusts health workers' information | 259 (92.2) | 22 (7.8) |

\*Combined "strongly disagree" and "disagree" with the negative statement

**Table 7.**
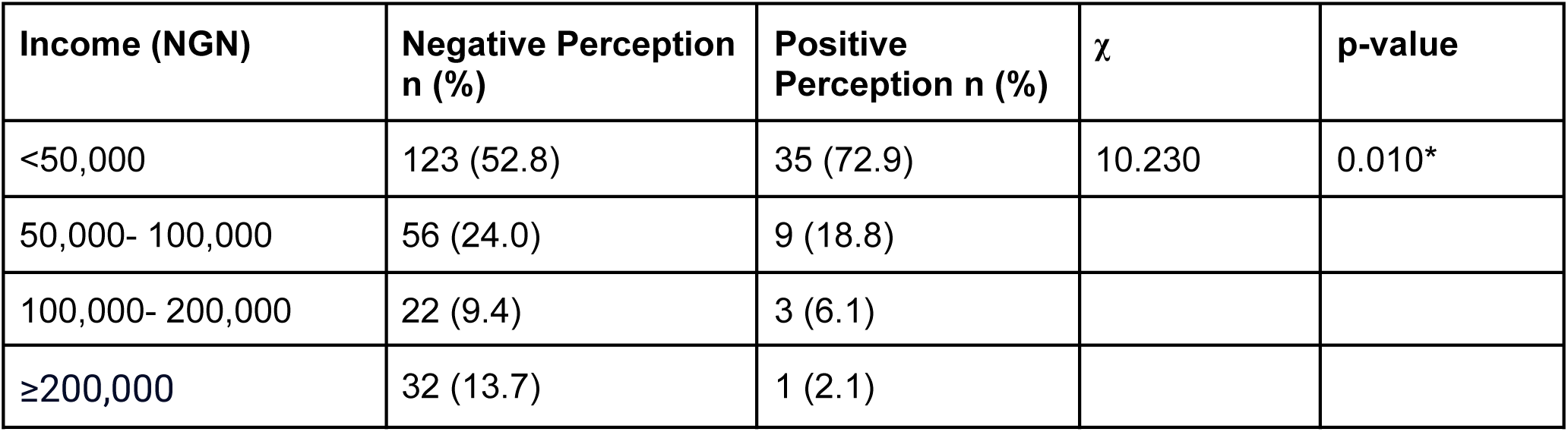
Association between household income and maternal perceptions of childhood vaccine safety (N = 281)

| Income (NGN) | Negative Perception n (%) | Positive Perception n (%) | $\chi^2$ | p-value |
| --- | --- | --- | --- | --- |
| <50,000 | 123 (52.8) | 35 (72.9) | 10.230 | 0.010* |
| 50,000- 100,000 | 56 (24.0) | 9 (18.8) |  |  |
| 100,000- 200,000 | 22 (9.4) | 3 (6.1) |  |  |
| ≥200,000 | 32 (13.7) | 1 (2.1) |  |  |

## Discussion

This study assessed the knowledge, attitudes, and perceptions (KAP) of mothers of under-five children attending primary healthcare centers in Abuja Municipal Area Council (AMAC), Nigeria, regarding routine childhood immunization. The findings reveal an overall satisfactory level of knowledge, generally positive attitudes, and favorable perceptions among respondents. The discussion highlights how sociodemographic factors intersect with immunization behaviors and how these findings align with or diverge from previous literature.

### Sociodemographic Characteristics

Most respondents were aged 25–34 years (61.9%), with a mean age of 28.8 years, consistent with similar studies (Abimbola *et al.,* 2023), which show that young mothers are the primary caregivers of under-five children. A large proportion (47.7%) had completed secondary education, and 38.4% had tertiary education, indicating a relatively educated cohort. The overwhelming majority (97.2%) were married, reflecting prevailing cultural norms, in line with reports by Adedire *et al*. (2021) and Abimbola *et al*. (2023). While 52.7% were employed, many earned below ₦50,000, possibly restricting healthcare access. The Igbo ethnic group was the most represented, echoing patterns reported by Galadima *et al*. (2021), where Igbo mothers were more likely to vaccinate their children than other ethnic groups. Cultural and religious influences were evident in shaping health behaviors and vaccination decisions, reflecting the multicultural nature of AMAC.

### Knowledge of Routine Childhood Immunization

Knowledge levels were high, with 94.7% of respondents acknowledging that vaccines protect against infectious diseases. These findings are consistent with global studies from Saudi Arabia (Almutairi *et al.,* 2021), Asia (Aung *et al.,* 2020), and Nigeria (Solomon *et al.,* 2024). However, notable knowledge gaps exist—37.4% were unaware of available vaccines, and 35.6% were unfamiliar with common side effects like fever. Alarmingly, 13.2% perceived vaccines as dangerous, aligning with trends of vaccine hesitancy influenced by misinformation (Marvila *et al.,* 2023; Mahachi *et al.,* 2022). While 90% correctly identified the need for vaccines at birth, only 76.9% appreciated the severity of vaccine-preventable diseases (VPDs), suggesting targeted health education is still needed.

### Attitudes Toward Immunization

Attitudes were predominantly positive. Nearly all respondents possessed vaccination cards (97.2%) and had vaccinated their children (96.8%), signifying strong adherence to national schedules. These findings align with those from the U.S. and Nigeria (Jayaraj *et al.,* 2023; Adedire *et al.,* 2021). However, uptake of supplementary vaccines (e.g., influenza, typhoid) was low (17.8%), a sharp contrast to 88.2% reported by Abimbola *et al*. (2023). Cost appears to be a major barrier, with 53% citing affordability concerns, consistent with reports from Jordan (Omayah *et al.,* 2023). Despite these barriers, 94.7% would recommend vaccines to others, underscoring broad public confidence.

### Perceptions of Immunization

Perceptions were largely favorable. More than half (55.2%) rejected the notion that vaccines are ineffective, although 38.8% remained unsure about vaccine potency. A strong majority (97.5%) believed vaccines are safe, echoing trust documented in earlier Nigerian studies (Adedire et al., 2021; Adeniyi *et al.,* 2023). Perceptions of government support were also positive, with 86.9% expressing satisfaction. However, 20.6% of mothers considered the religion of healthcare workers when deciding to vaccinate, pointing to underlying sociocultural influences requiring further community-level investigation.

### Sociodemographic Associations with Knowledge

Educational attainment significantly influenced knowledge levels (p < 0.001). Mothers with secondary or tertiary education had superior knowledge, aligning with findings from Georgia, India, Ethiopia, and Nigeria (Tengiz *et al.,* 2019; Jayaraj *et al.,* 2023; GebreEyesus *et al.,* 2021). Similarly, household income showed a strong positive association with knowledge (p < 0.001), affirming that economic capacity enhances access to health information. Other variables—including age, marital status, religion, and ethnicity—did not significantly affect knowledge levels, suggesting education and income are the dominant drivers.

### Sociodemographic Associations with Attitudes

No demographic factor was significantly associated with attitudes, although variation existed. For instance, employed mothers and those with secondary education displayed slightly more positive attitudes, aligning with Budu *et al*. (2023). However, this trend did not reach statistical significance, suggesting that while economic and educational empowerment may influence attitudes, other psychosocial or contextual factors may also play roles.

### Sociodemographic Associations with Perceptions

Only household income showed a statistically significant association with perception (p = 0.010). Interestingly, mothers from lower-income brackets showed more favorable perceptions. This finding may reflect greater reliance on public health systems and exposure to government outreach efforts. It contrasts with other studies (Cooper *et al.,* 2021; Fenta *et al.,* 2021; Budu *et al.,* 2023) that associate higher income with better vaccination outcomes. Other demographic factors did not significantly affect perception.

### Determinants of Knowledge

Logistic regression analysis confirmed that higher income significantly predicted good knowledge of immunization (aOR = 6.51, p = 0.007). This aligns with studies from Saudi Arabia (Almutairi *et al.,* 2021), Asia (Aung *et al.,* 2020), and Pakistan (Shahzadi *et al.,* 2022), which emphasize the central role of economic empowerment in improving health literacy and immunization uptake.

### Determinants of Attitude

No demographic factors significantly predicted attitudes toward immunization in regression analysis. However, trends suggest that employed and higher-income mothers might exhibit more favorable attitudes, albeit not statistically significant. These trends suggest that broader structural factors, such as access, convenience, and public messaging, may be more influential than demographic traits.

### Determinants of Perception

Perception was significantly influenced by income level. Mothers earning between ₦100,000–₦200,000 were less likely to have positive perceptions than those earning below ₦50,000 (aOR = 0.09, p = 0.025). This surprising outcome may reflect the competing demands of working mothers, as previously suggested by Danso *et al*. (2023). Other variables—including age, education, marital status, employment, and religion—showed no significant effect, reinforcing the dominant influence of economic context on perception.

## Conclusion

This study assessed the knowledge, attitudes, and perceptions of mothers of under-five children attending primary healthcare centers in Abuja Municipal Area Council (AMAC) regarding routine childhood immunization. The findings reveal that, although general awareness and attitudes toward immunization were positive, significant knowledge gaps persist, especially concerning specific vaccines and their possible side effects. Socioeconomic factors such as education level and household income played a critical role in shaping mothers’ knowledge and perceptions, whereas demographic factors like age and marital status showed minimal impact. A major barrier identified was the financial constraint affecting the uptake of supplementary vaccines.

Overall, the study highlights the urgent need for targeted, multifaceted interventions to improve maternal understanding, overcome financial obstacles, and address sociocultural factors that influence vaccine acceptance and uptake in the AMAC region.

## Data Availability

All data produced in the present study are available upon request to the author

## Acknowledgements

I would like to thank the study participants for their openness and enthusiasm in participating in this study. I would also like to express our immense gratitude to Dr. Darma Abdu Mohammed for his support in guiding me through the project. I would also like to thank the entire Primary Health Care Centers within Amac Abuja for their collective support and encouragement throughout the study process.

